# Integrating Prehabilitation into Surgical Pathways: Current Modalities, Outcomes, and Research Gaps

**DOI:** 10.1101/2025.09.23.25336233

**Authors:** Eric Sosa, Anabel Henick, Dhanesh D. Binda, Crystal Joseph, Stanley Kim, Dave Mathew, Singh Nair, Jinu Kim, David C. Adams, Karina Gritsenko, Naum Shaparin

## Abstract

**Background:** Prehabilitation constitutes a multidisciplinary strategy aimed at optimizing patients’ physiological and psychological status prior to surgery, with the objective of improving postoperative outcomes and long-term quality of life. Interventions commonly include structured exercise programs, nutritional optimization, respiratory muscle training, and psychosocial support. This narrative review synthesizes the expanding body of evidence on prehabilitation, with particular emphasis on the implementation of diverse modalities, their impact on surgical outcomes, and the persisting gaps in knowledge that warrant further investigation.

**Methods:** A systematic PubMed search was performed on June 25, 2025, using the terms “Prehabilitation AND Anesthesia,” “Prehabilitation AND Surgery,” “Prehabilitation AND Preoperative Medicine,” and “Prehabilitation.” Extracted data included article title, year of publication, surgical specialty, procedure type, study design, prehabilitation modality, number of modalities, regimen duration, demographic characteristics, loss to follow-up, cancer status or receipt of neoadjuvant chemotherapy, study outcomes, data collection methods, and study conclusions. Systematic reviews, meta-analyses, protocol descriptions, and feasibility studies without reported results were excluded from the final analysis.

**Results:** The search strategy identified 796 articles, 76% of which were published between 2017 and 2025. A total of 153 studies met inclusion criteria for analysis. The most frequently represented surgical specialties were general surgery (43%), orthopedic surgery (21%), and cardiothoracic surgery (18%). Single-modality prehabilitation was reported in 43% of studies, whereas multimodal approaches varied by specialty, occurring in 36% of general surgery studies and 13% of cardiothoracic surgery studies. Five principal prehabilitation modalities were identified: exercise, nutrition, psychological intervention, substance cessation, and medical optimization. Exercise-based interventions were the most common, incorporated in 84.7% of studies, followed by nutritional interventions in 29.5%. Overall, 82% of included studies reported statistically significant improvements in surgical outcomes associated with prehabilitation.

**Conclusion:** Most studies included in this review demonstrated a significant positive impact of prehabilitation on surgical outcomes. Nonetheless, most investigations employed a single prehabilitation modality, underscoring the need to further assess the effectiveness of multimodal strategies. Furthermore, the existing evidence is concentrated within a limited number of surgical specialties, highlighting the necessity for well-designed studies across a broader spectrum of surgical disciplines.

## Introduction

Population projections indicate that the number of individuals aged 65 years and older will double within the next decade.^1^ In conjunction with age-related physiological decline and the increased prevalence of frailty, this demographic shift is expected to impose substantial strain on perioperative services, heighten demand for specialized geriatric care, and underscore the importance of strategies such as prehabilitation to optimize outcomes in this vulnerable population.^2,3^

The physiologic stress response to surgery in deconditioned, older, and frail patients is strongly associated with increased postoperative morbidity and mortality.^4^ Adverse surgical outcomes in both the short and long term demonstrate a proportional relationship with the burden of comorbid conditions, including postoperative infections, impaired wound healing, pulmonary complications, cardiovascular events, renal dysfunction, prolonged hospitalization, readmissions, and increased mortality.^5–7^ These risks are particularly pronounced in elderly patients, who frequently present with multiple comorbidities that contribute to frailty, such as cardiovascular disease, chronic obstructive pulmonary disease, diabetes mellitus, chronic kidney disease, malnutrition, and cognitive impairment.^8^ Although advances in perioperative medicine have improved outcomes in this population, a substantial risk of complications persists, shaped by the interplay of physical, psychological, and social determinants of health. Traditionally, postoperative rehabilitation has been central to surgical recovery; however, its overall effectiveness has been increasingly called into question.^9–15^

The concept of “prehabilitation” was first introduced in 1946 by the British military to describe interventions intended to correct physical deficits prior to fitness evaluations.^16^ Scientific interest in prehabilitation reemerged in the 1980s, primarily within the domain of sports medicine.^17^ In contemporary perioperative medicine, prehabilitation is defined as the optimization of a patient’s preoperative status with the goal of reducing postoperative complications and improving quality of life.^18^ Interventions commonly include structured aerobic and resistance exercise programs, nutritional optimization (e.g., protein supplementation), respiratory training such as inspiratory muscle exercises, and psychosocial support including stress reduction and counseling. Poor preoperative physical condition has consistently been linked to higher rates of postoperative morbidity and mortality, and prehabilitation strategies are intended to mitigate these risks.^19–21^

Recent prehabilitation research has predominantly focused on colorectal surgery.^22–25^ Positive findings from these studies have spurred interest in exploring prehabilitation in additional surgical specialties, including urology and gynecology.^26,27^ Overall, current evidence demonstrates that prehabilitation enhances physical function, reduces postoperative complications, and shortens hospital length of stay.^28,29^

Despite growing evidence supporting the safety and efficacy of prehabilitation, it has not yet been widely adopted as a standard of care, and the existing body of literature remains limited.^30^ This comprehensive review is therefore both timely and necessary, providing a critical examination of the evolution and expansion of prehabilitation across surgical specialties, the diversity and duration of its modalities, and its measurable impact on surgical outcomes. The review synthesizes current evidence and identifies key gaps in knowledge to guide future research directions and support the integration of prehabilitation into routine preoperative care, with the goal of enhancing patient resilience, optimizing recovery, and improving long-term outcomes.

## Methods

### Search Strategy

A structured and reproducible search strategy was developed, informed by methodologies established in prior systematic reviews. The search was performed on June 25, 2025, using the PubMed database and was designed to identify literature specific to prehabilitation in the surgical setting. Search terms included “prehabilitation AND anesthesia,” “prehabilitation AND surgery,” “prehabilitation AND preoperative medicine,” and “prehabilitation.” The Boolean operator “AND” was applied to refine results and ensure relevance to preoperative care, while the strategy was deliberately broad to maximize sensitivity and capture the widest possible range of eligible studies.

### Study Selection

Two reviewers independently screened articles tagged with the specified search terms to identify studies relevant to prehabilitation. Eligible studies included clinical trials, both randomized and non-randomized, as well as retrospective analyses. Studies examining the Enhanced Recovery After Surgery (ERAS) protocol, prehabilitation combined with rehabilitation, and feasibility studies with reported outcomes were also included. Exclusion criteria encompassed systematic reviews, meta-analyses, general reviews, protocol descriptions, case reports, opinion pieces, and letters to the editor. Duplicate entries across the four searches were removed.

### Data Extraction

For all included articles in the analysis, the following data were extracted: PubMed ID, article title, year of publication, surgical specialty, type of surgery, study design, prehabilitation modality or modalities, inclusion of cancer patients, receipt of neoadjuvant chemotherapy, sample size, mean age, gender distribution, duration of prehabilitation, involvement of ERAS protocols, number lost to follow-up, primary and secondary outcomes, data collection methods, and study conclusions. Cardiothoracic surgeries were subcategorized into two groups: Group 1A included true cardiac procedures (e.g., coronary artery bypass grafting and valve replacements), while Group 1B encompassed oncologic procedures (e.g., esophagectomies and pulmonary resections). Cancer or neoadjuvant chemotherapy status was recorded only when explicitly stated in the study. Study conclusions were classified as “positive,” “negative,” or “equivocal” based on the statistical significance and overall interpretation of findings reported in the articles. No additional statistical analyses were performed beyond those presented in the source studies. All data are reported as counts and proportions, and analysis was conducted using Microsoft Excel Version 16.46. For interpretative clarity, studies classified as having “negative” or “equivocal” conclusions were grouped together, as neither reflected a clearly positive outcome.

### Categories of Prehabilitation

Prehabilitation regimens were classified into five primary categories: exercise, nutritional supplementation, psychological support, substance cessation, and medical optimization. The exercise category encompasses all forms of physical activity or training, including physical therapy, aerobic exercise, resistance or strength training, stretching, and walking. For urological studies, continence and pelvic floor training were also included within this category. Nutritional prehabilitation involved interventions such as modified dietary regimens, nutrition education, or consultations with a registered dietitian. Psychological support interventions included both in-person and virtual therapy sessions, emotional support groups, motivational interviewing, and anxiety management strategies. Substance cessation encompasses counseling aimed at discontinuing the use of alcohol, tobacco, and opioids. Medical optimization refers to interventions such as preoperative antibiotic regimens, improvement of glycemic control (e.g., reduction in glycosylated hemoglobin), anemia correction, and frailty management. While neoadjuvant chemotherapy was not considered a form of medical optimization in this classification, its inclusion was recorded during data collection due to its frequent occurrence in prehabilitation studies. The number of prehabilitation modalities used in each study was determined by the number of distinct prehabilitation categories represented in the intervention. For instance, a regimen that incorporated both general and pelvic floor exercises, along with regular follow-up sessions with a nutritionist, was classified as including two modalities: exercise and nutrition. Regimens incorporating two or more distinct categories of prehabilitation, such as exercise combined with nutritional support or psychological counseling, were classified as “multimodal.” Multimodal prehabilitation refers to the integration of multiple complementary interventions designed to address various physiological and psychosocial domains, with the aim of enhancing overall surgical readiness and recovery.

## Results

### Types of Studies Analyzed

The preliminary screening process identified 177 full-text articles for evaluation. Following full-text review, 24 articles were excluded due to irrelevance or lack of reported results, yielding a final cohort of 153 articles for comprehensive analysis (**Figure 1**). **Supplementary Table 1** details the characteristics of the included studies. Of the 153 studies analyzed, 45% were randomized controlled trials, 37% were prospective non-randomized cohort studies, 10% were retrospective reviews, and 5% were categorized as other study designs.

**Figure 1.**
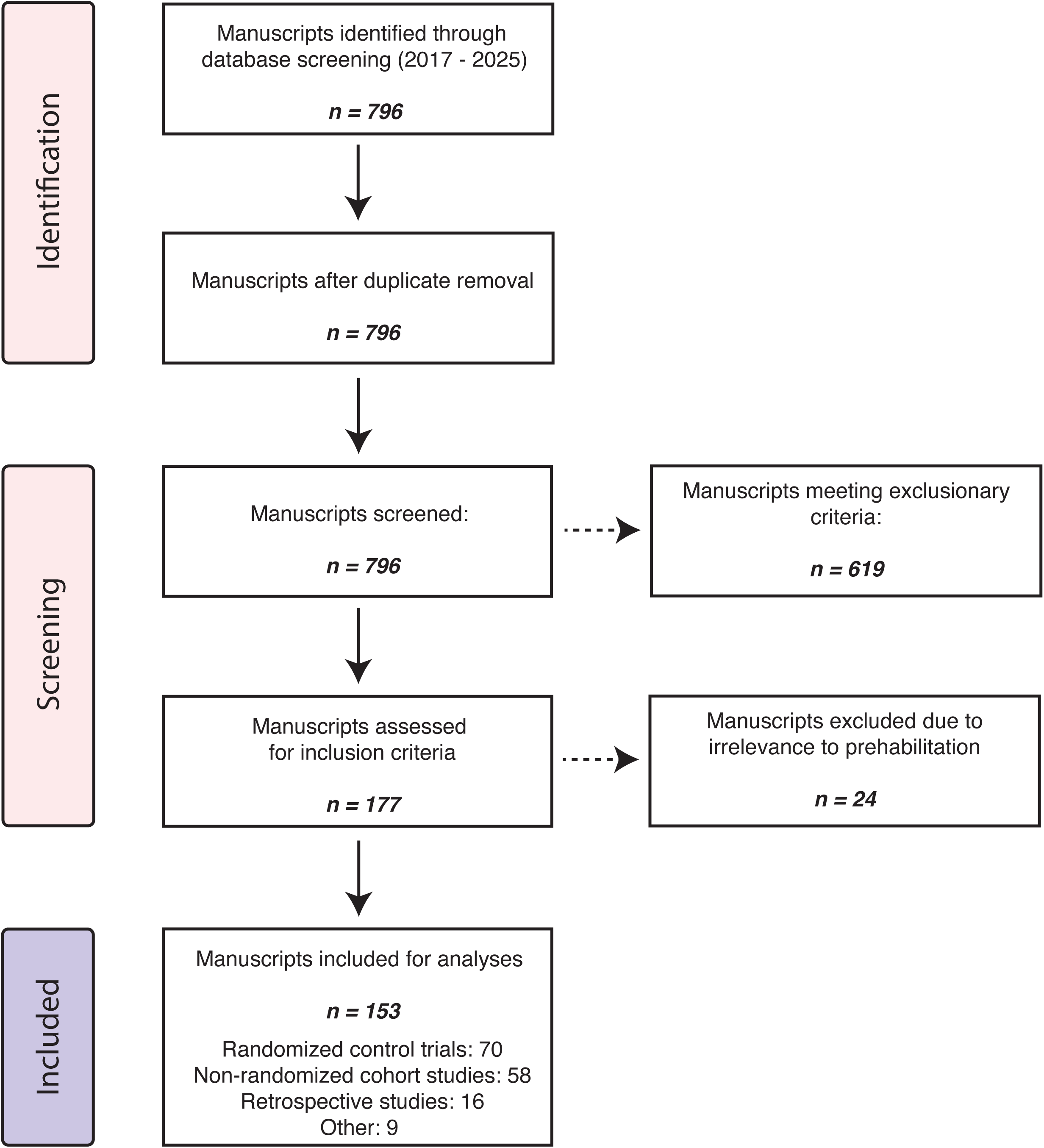
Study selection for analysis of prehabilitation modalities. Manuscripts published between 2015 and 2017 were screened. After de-duplication, 796 records remained; 619 were excluded as systematic reviews, meta-analyses, general reviews, protocol descriptions, case reports, opinion pieces, or letters to the editor. The remaining 177 full texts were assessed for eligibility, and 24 were excluded as not relevant to prehabilitation modality studies, yielding 153 studies for analysis: 70 randomized controlled trials, 58 non-randomized cohort studies, 16 retrospective studies, and 9 classified as other designs.

### Publication Timeline

Of the 153 studies analyzed, 124 (81%) were published between 2017 and 2020, and 29 (19%) between 2021 and 2025. The earliest clinical trial identified under the defined search criteria was published in 2008. Thereafter, the volume of publications increased markedly, with 12 articles appearing between 2009 and 2012, 17 between 2013 and 2016, and 123 between 2017 and 2025 (**Figure 2A**), reflecting the growing academic interest in prehabilitation.

**Figure 2.**
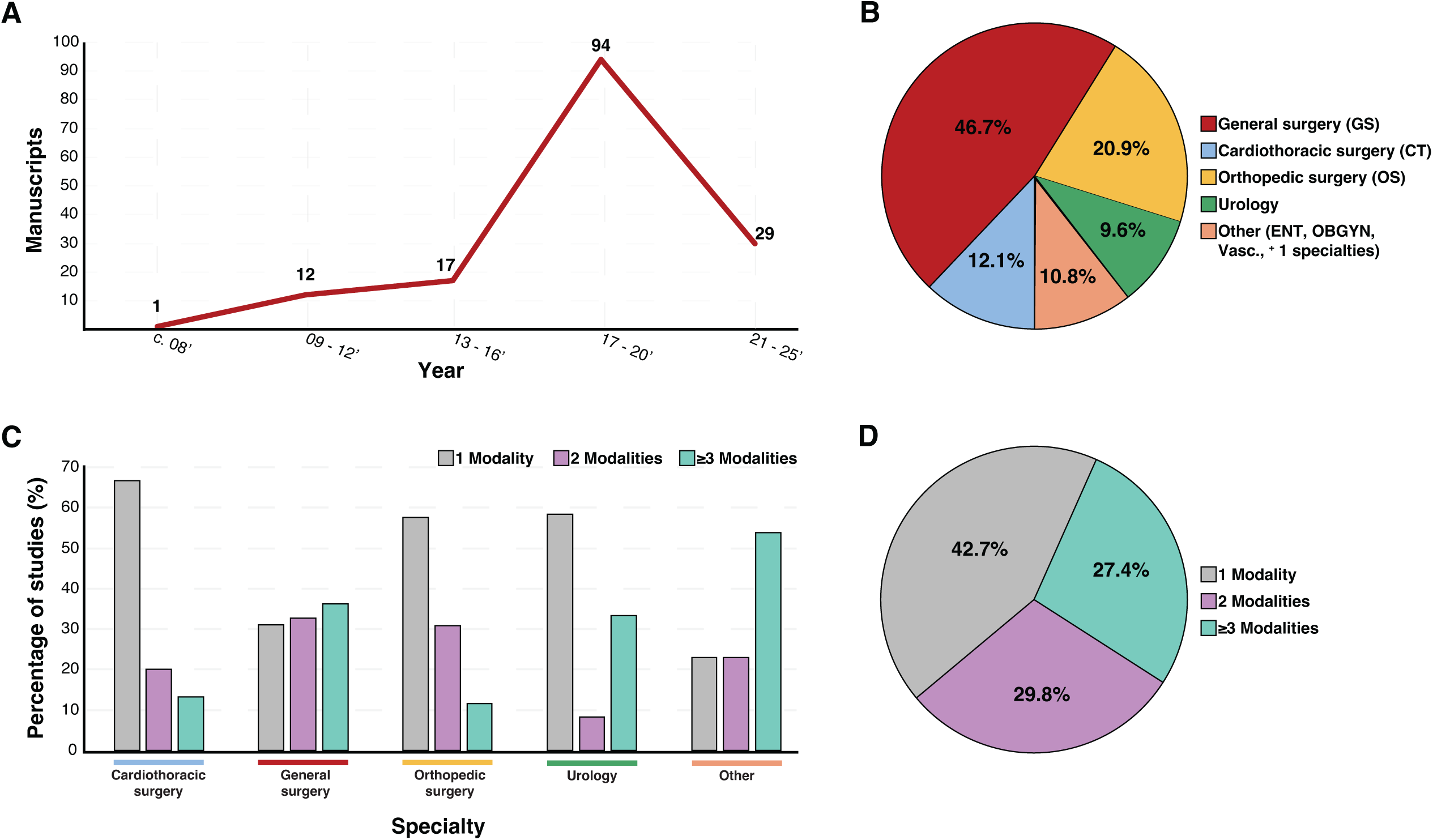
Temporal trends, specialty distribution, and modality composition of prehabilitation manuscripts. *A*. Prehabilitation manuscripts between 2008 and 2025 (x-axis) indicate an increase in prehabilitation modality usage (y-axis). The x-axis representing the year of publication was divided into three-year increments. In 2008, there was 1 publication, increasing to 12 between 2009–2012, 17 between 2013–2016, and a surge between 2017–2020 with 94 manuscripts applying prehabilitation modalities. ***B***. Pie chart highlighting the specialty composition of the 153 studies analyzed. General surgery (red) comprised the greatest proportion at 46.7%, followed by orthopedic surgery at 20.9% (yellow), cardiothoracic surgery at 10.8% (blue), urology at 9.6% (green), and the Other category (ENT, obstetrics/gynecology, vascular, and multispecialty) at 12.1% (tan). The “Other” category included studies spanning more than one specialty. ***C***. Representation of the number of prehabilitation modalities employed in the analyzed studies by specialty. The use of one, two, or three or more prehabilitation modalities is represented by grey, purple, and blue bars, respectively. The percentage of studies employing these modalities is shown on the y-axis. ***D***. Pie chart visual representation of the number of modalities used in the analyzed studies in aggregate. 42.7% of studies used one modality, 29.8% used two, and 27.4% utilized three or more modalities.

### Demographic Characteristics

The mean patient age across all studies was 62.4 years (SD = 11.6), with reported ages ranging from 23 to 80.6 years. Age varied by surgical specialty: orthopedic surgery studies reported the youngest average age (60.3 years; SD = 14.7), while urology studies had the oldest (65.8 years; SD = 4.7). Cardiothoracic and general surgery studies reported mean ages of 64.0 years (SD = 5.8) and 61.8 years (SD = 12.8), respectively. The mean sample size was 130.9 patients (SD = 230.8), with considerable variability due to differences in study design. Sample sizes ranged from 6 (one-arm intervention trial) to 2,187 (retrospective review). Variability in sample size also differed by specialty, with orthopedic surgery studies demonstrating the largest standard deviation (SD = 351.3) and cardiothoracic surgery the smallest (SD = 85.7). Gender distribution favored male patients, with an overall male-to-female ratio of 1.41:1. Cardiothoracic surgery studies had the highest male predominance (2.1:1), while orthopedic surgery had the lowest (0.8:1). Loss to follow-up was reported in 96 of the 153 studies. On average, 85.2% (SD = 14.7%) of participants completed follow-up. Cardiothoracic surgery studies had the highest follow-up completion rate (mean = 89.1%; SD = 11.2%), whereas studies involving multiple surgical specialties had the lowest (mean = 81.0%; SD = 21.8%). The average duration of prehabilitation programs was 5.8 weeks (SD = 4.7), with variability across studies.

### Oncologic Context

Most studies (56.3%) focused on prehabilitation in cancer patients undergoing surgery. However, only 23.5% included patients receiving neoadjuvant chemotherapy. Orthopedic surgery studies did not include cancer patients or those undergoing neoadjuvant therapy. In contrast, urology studies had the highest proportion of cancer-focused prehabilitation (80%), predominantly in the context of prostate and bladder cancer resections.

### Surgical Specialties

The most frequently represented surgical specialties among the 153 analyzed studies were general surgery (46.7%), orthopedic surgery (20.9%), cardiothoracic surgery (12.1%), and urology (9.6%) (**Figure 2B**) (**Table 1**). Specialties with two or fewer studies—such as otolaryngology, vascular surgery, and obstetrics and gynecology—as well as studies involving multiple surgical disciplines (approximately 10.8%) were collectively categorized as “Other Studies” for the purpose of analysis.

Within general surgery, colorectal procedures accounted for most studies (47%). In orthopedic surgery, prehabilitation was most investigated in joint replacement (68%) and spine surgeries (21%). For cardiothoracic surgery, pulmonary resections (42%) and esophageal resections (24%) were the predominant focus. In urology, prostatectomies (61%) and cystectomies (33%) represented the most studied procedures. Additional surgeries included in the dataset, though less frequently represented, involved organ transplantation, coronary artery bypass grafting, hepatopancreatic biliary procedures, and hernia repairs.

### Patterns of Prehabilitation Modality Use Across Surgical Specialties

Studies were further analyzed for the number of prehabilitation modalities employed by surgical specialty (**Figure 2C**). In cardiothoracic surgery (*blue line*), 67% of studies used a single modality, with 20% and 15% employing two and ≥3 modalities, respectively. In general surgery (*red line*), the number of prehabilitation modalities employed was more evenly distributed across single, two, and ≥3 modalities (31%, 33%, and 37%, respectively), indicating a greater uptake of multimodal programs than single modality alone. Orthopedic surgery (*yellow line*) and urology (*green line*) predominantly used a single modality (≈60% in each), with smaller shares employing two or ≥3 modalities. The pooled “other” specialties (orange line)—otolaryngology, gynecology, vascular surgery, and smaller fields—were the only subgroup in which ≥3 modalities predominated (54%), exceeding the proportions using one or two modalities (22% and 22%, respectively). Across the cohort, single-modality studies accounted for 42.7%, two-modality for 27.4%, and ≥3-modality for 29.8% of all studies (**Figure 2D**). These findings underscore the predominance of single-modality programs while highlighting a meaningful—though still limited—shift toward multimodal prehabilitation strategies.

### Types and Prevalence of Prehabilitation Modalities

Five distinct categories of prehabilitation were identified: exercise, nutritional support, psychological interventions, substance cessation, and medical optimization. Exercise was the most frequently employed modality, present in 87.1% of studies, followed by nutritional interventions (37.9%), psychological support (32.6%), medical optimization (11.2%), and substance cessation (11.2%) (**Figure 3A**). Of the 153 studies, 59% (n = 90) utilized a single prehabilitation modality (**Table 1**). Multimodal prehabilitation—defined as the use of two or more distinct intervention categories—was employed in 20% of studies (n = 31), though implementation varied by specialty. General surgery had the highest proportion of multimodal regimens (27%), while cardiothoracic surgery had the lowest (10%) (**Figure 3B**). The overall mean number of prehabilitation modalities used per study was 1.98. This also varied by specialty, with general surgery reporting the highest average number of modalities (2.12), and cardiothoracic surgery the lowest (1.46).

**Figure 3.**
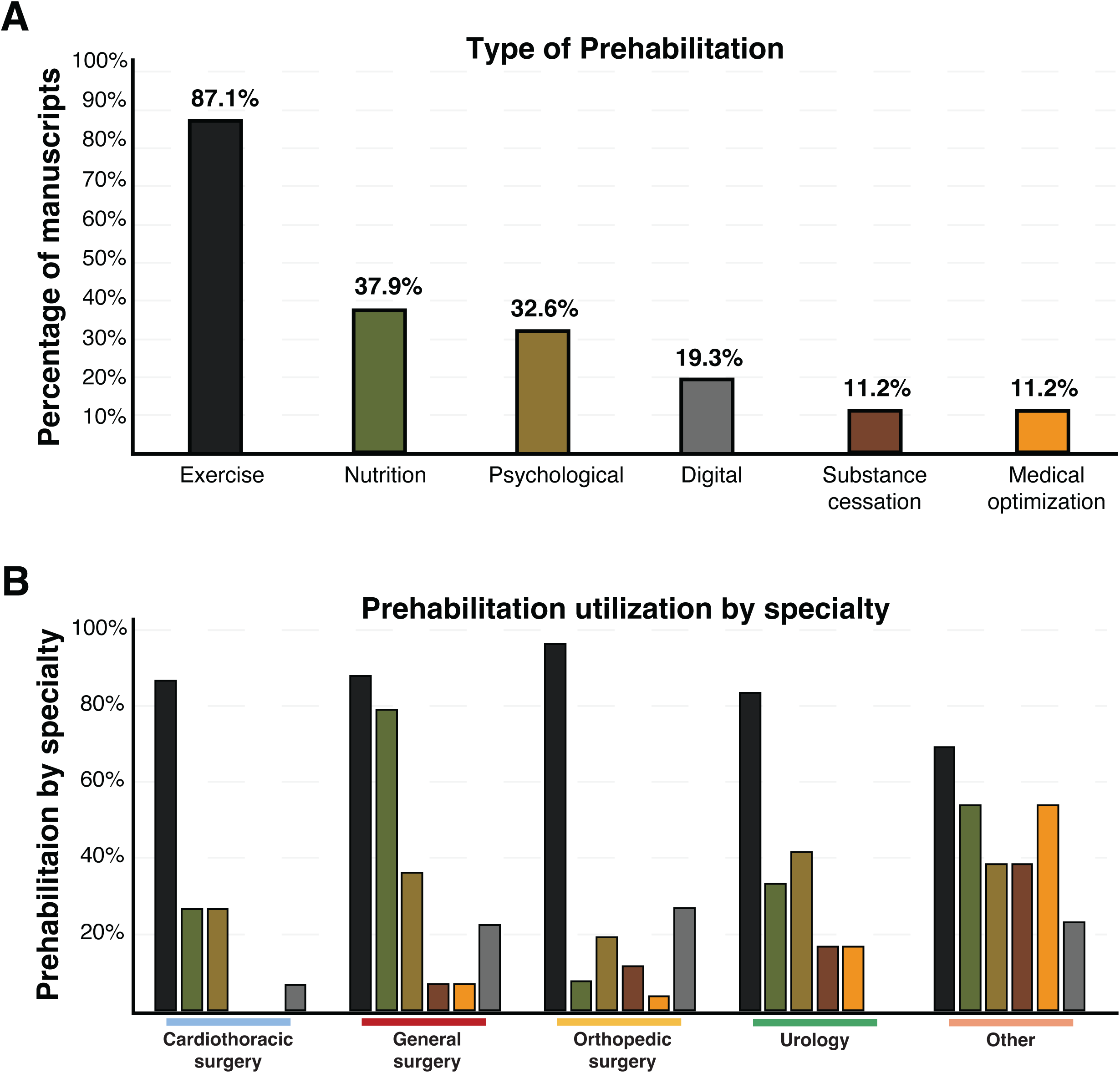
Distribution of prehabilitation modalities and utilization by surgical specialty. ***A***. Percentage of manuscripts that reported each prehabilitation modality (y-axis); bars were color-coded according to the following scheme: exercise (black, 87.1%), nutrition (green, 37.9%), psychological (light brown, 32.6%), digital (grey, 19.3%), substance cessation (dark brown, 11.2%), and medical optimization (orange, 11.2%); modalities were ordered by frequency, with exercise the most utilized. ***B***. Prehabilitation utilization subdivided by surgical specialty (x-axis: cardiothoracic surgery, general surgery, orthopedic surgery, urology, other) with the percentage of studies on the y-axis; bars were colored by modality according to the scheme in A and applied consistently across the panel.

### Proportion of Positive Conclusions

Across all included studies, 82% reported a “positive” conclusion, indicating that prehabilitation produced a statistically significant improvement in either primary or secondary outcomes, as defined by the original study authors (**Figure 4**). The proportion of positive conclusions varied by surgical specialty. General surgery (*red bar*) had the highest rate of positive findings at 93%, followed by urology (*green bar*) at 83%, cardiothoracic surgery (*blue bar*) at 80%, and orthopedic surgery (*yellow bar*) at 73% (**Table 1**).

**Figure 4.**
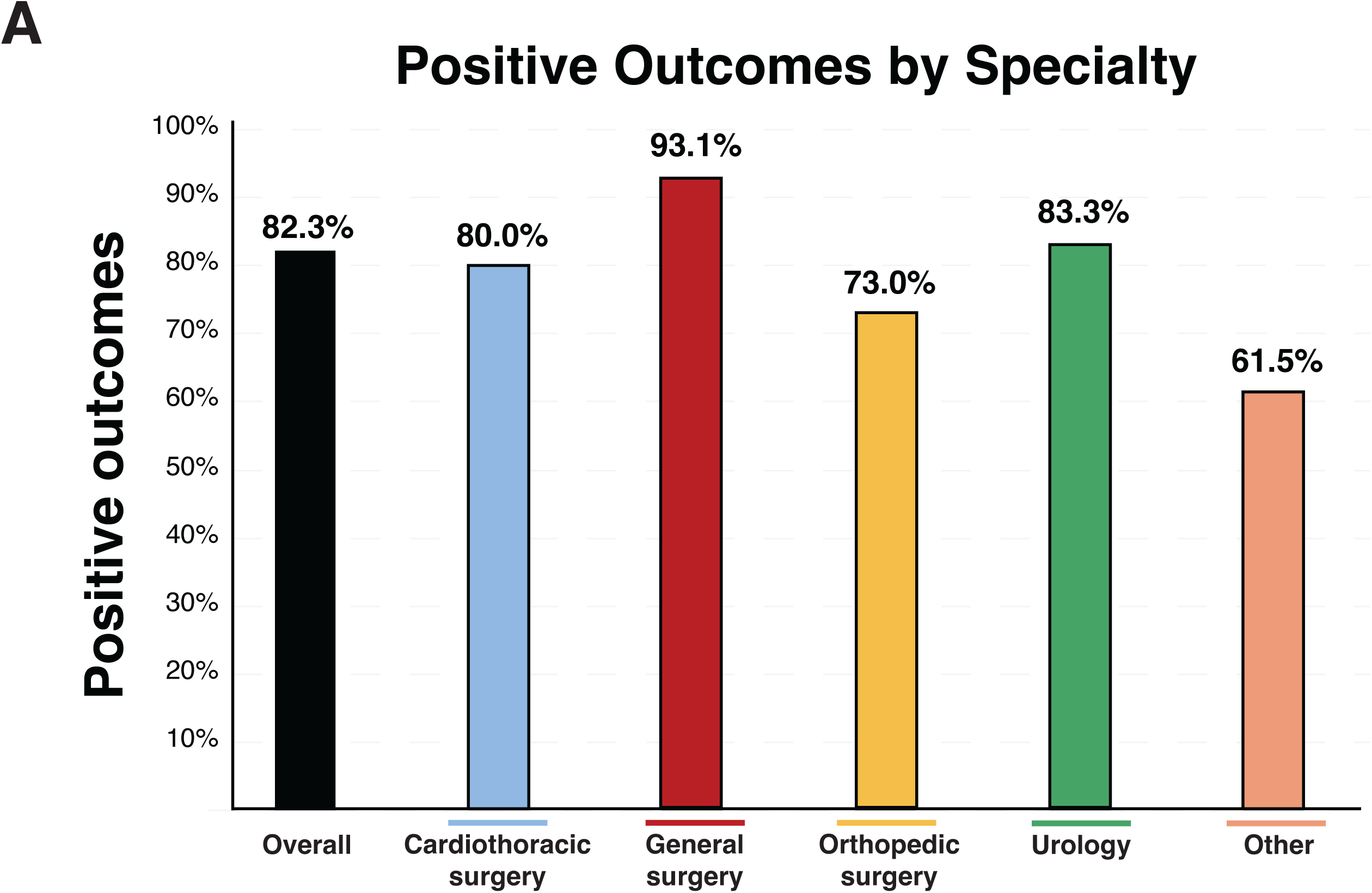
Positive outcomes associated with prehabilitation by surgical specialty. ***A*.** Proportion of included studies reporting positive outcomes (y-axis, %) presented overall and stratified by specialty (x-axis). Positive outcomes were observed in 82.3% overall; by specialty: cardiothoracic surgery, 80.0%; general surgery, 93.1%; orthopedic surgery, 73.0%; urology, 83.3%; and other, 61.5%. Percentages were calculated from the included dataset (n = 153); “other” comprised ENT, obstetrics/gynecology, vascular, and multispecialty studies.

## Discussion

Since 2008, research on prehabilitation has expanded rapidly, with most studies in our review published between 2017 and 2025, underscoring prehabilitation as a relatively recent focus in perioperative care. Analysis of demographic data revealed a predominance of male participants across studies, with an overall male-to-female ratio of 1.4:1. This contrasts with findings from a previously published review of seven studies, in which five demonstrated gender parity and two showed a female predominance.^31^ The most pronounced gender disparity was observed in cardiothoracic surgery studies, where the male-to-female ratio reached 2.1:1. This likely reflects the higher incidence of lung cancer in men, as pulmonary resections—commonly indicated for lung malignancies—represented the most frequently studied procedure within this specialty.^32^ The mean age of participants was in the elderly range, aligning with the increased surgical risk, frailty, and comorbidity burden typically observed in older populations.^32–34^ Given that a majority of studies reporting positive outcomes involved elderly and predominantly male patients, these populations may stand to derive the greatest benefit from prehabilitation interventions.

### Adherence and Patient Engagement

Across all studies analyzed, the average rate of adherence at follow-up was 85.2%. Notably, cardiothoracic surgery studies reported the highest adherence rates. This is particularly compelling given that cardiac surgery patients typically present with multiple comorbidities, which are known to contribute to higher perioperative risk and potential loss to follow-up.^25^ Several factors may explain this elevated adherence. Patients undergoing cardiothoracic procedures, such as coronary artery bypass grafting (CABG) or valve replacement, may have a heightened awareness of the risks associated with their condition and the complexity of postoperative recovery. This awareness, possibly driven by fear or a clearer understanding of the severity of their disease, may enhance motivation to engage with prehabilitation and follow-up protocols. Additionally, evidence suggests that prehabilitation in cardiac patients yields substantial benefits. In a recent observational study evaluating prehabilitation prior to CABG, participants demonstrated significant improvements in cardiorespiratory fitness, body mass index, and psychological outcomes, such as symptoms of depression and anxiety, alongside increased participation in postoperative cardiac rehabilitation programs.^26^ These findings indicate that prehabilitation not only improves measurable health outcomes but may also foster sustained patient engagement in long-term health optimization strategies.

### Prevalence of Prehabilitation Across Surgical Specialties

From our analysis, the primary surgical specialties utilizing prehabilitation, in order of frequency, were general surgery, orthopedics, cardiothoracic surgery, and urology. Previous systematic reviews have demonstrated similar distributions.^35–37^ Across the literature, five core prehabilitation modalities were identified: exercise, nutritional planning, psychological intervention, substance cessation, and medical optimization.^38,39^ Despite this breadth, most studies implemented only a single modality, with exercise being the most frequently employed. The mean number of modalities per study was 1.7, which likely reflects the disproportionate impact of the relatively small number of studies that incorporated several interventions, thereby raising the average above one. Given the evidence that multimodal prehabilitation can reduce morbidity and mortality,^40,41^ this represents a critical opportunity for improvement. Additional research is warranted to assess the efficacy of multimodal interventions. By incrementally integrating multiple components, prehabilitation could be delivered in a comprehensive, single-visit framework, yielding a more holistic, efficient, and clinically effective approach.

### Targeted Subspecialty Approaches

In some subspecialties, prehabilitation modalities were highly targeted. For example, several urological studies employed pelvic floor muscle training to mitigate post-procedural urinary incontinence,^42,43^ while select otolaryngological studies administered gentamicin to accelerate vestibular function recovery.^44,45^ These examples illustrate how refined, indication-specific prehabilitation strategies may prevent common complications in defined patient subsets. Future research aimed at identifying prevalent complications and matching them with targeted interventions could inform best practices for subspecialty-specific prehabilitation.

### Clinical Outcomes and Knowledge Gaps

The literature predominantly demonstrates positive postoperative outcomes associated with prehabilitation. Improvements in functional capacity and quality of life were consistently reported in general surgery, cardiothoracic surgery, and urology.^28,30^ Despite these encouraging results, important knowledge gaps remain. As a relatively novel area of investigation, additional studies are needed to further delineate the role of prehabilitation within the framework of ERAS pathways, which have achieved widespread adoption across most surgical specialties.^43,46^

### Underutilized Specialties and Disparities

Several surgical subspecialties, including neurosurgery, otolaryngology, and vascular surgery, rarely employed prehabilitation. The limited uptake in these fields represents another opportunity for exploration. The reasons underlying this underutilization remain unclear but may include patient-level barriers or the urgency associated with operative intervention. Additionally, demographic disparities were evident, with younger patients and female patients underrepresented in published cohorts. Addressing these gaps in future research will be essential to ensure prehabilitation protocols are representative of the broader surgical population.

### Risk Stratification and Long-Term Outcomes

Incorporating surgical risk stratification into prehabilitation studies represents another potential area of advancement.^44^ Many studies combined heterogeneous surgical practices, complicating efforts to determine whether procedural risk influenced prehabilitation outcomes. Stratification by procedural invasiveness could better clarify the relationship between prehabilitation and clinical endpoints. Furthermore, while most studies focused on short-term outcomes (<3 months), the long-term durability of prehabilitation benefits remains poorly characterized. Investigation into long-term outcomes will be critical to defining its sustained clinical value.

### Economic Implications

Evidence regarding the financial implications of prehabilitation is limited and inconsistent. Few studies have directly assessed its cost-effectiveness, as interventions are often initiated exclusively within research contexts. Since prehabilitation is not yet a standard of care, it is not routinely prescribed, and therefore, limited guidance exists regarding its cost–benefit balance. While some studies have reported cost savings,^47,48^ others have demonstrated neutral or unfavorable findings.^49^ For example, a 2019 cost-analysis reported no significant savings at 30 days among patients enrolled in a prehabilitation program.^49^ Further research is required to elucidate the economic impact of prehabilitation, particularly in real-world settings.

### Digital Delivery Models

Implementation of prehabilitation also entails considerable resource demands and patient time commitments. Digital modalities were not classified as independent prehabilitation modalities in this review, owing to their limited use and overlap with other interventions, most commonly exercise. ^45^ Nonetheless, digital delivery represents a promising means of improving accessibility and reducing costs. Patients with mobility or transportation barriers may particularly benefit from tele-prehabilitation, which could extend care directly to patients’ homes. Evaluation of digital and hybrid models is warranted to assess feasibility, accessibility, and clinical effectiveness.

### Social Determinants of Health

Social determinants of health are another underexplored factor in prehabilitation implementation. Employment status, income, health literacy, and insurance coverage may all serve as barriers to participation, particularly when programs are costly, lengthy, or rigidly scheduled. Future investigations should examine how these determinants influence access, adherence, and outcomes, and design strategies to mitigate disparities.

### Publication Bias

Adoption of prehabilitation into routine surgical care is also influenced by the integrity and balance of the scientific literature. Publication bias is well recognized in clinical research, and although the extent of this bias in prehabilitation studies has not been quantified, the predominance of positive findings in the published literature is notable. Future investigations should assess the degree of publication bias to ensure balanced evidence synthesis.

### Clinical Integration

Elective surgery provides a unique opportunity for addressing uncontrolled or poorly managed chronic conditions through multidisciplinary collaboration and preoperative care coordination. Individualized medical optimization, including prehabilitation, has the potential to reduce preventable postoperative complications and should be considered for integration into standard preoperative planning. While the cost-effectiveness of such programs remains incompletely defined, efforts to implement affordable, accessible, and evidence-based prehabilitation protocols may serve to reduce disparities in surgical outcomes.

## Limitations and Future Directions

Several limitations of our review should be acknowledged. Categorization of prehabilitation modalities was based on published studies; however, interventions such as substance cessation and medical optimization may be more accurately considered elements of routine preoperative care rather than distinct prehabilitation modalities. Similarly, digital prehabilitation may be best classified as a method of delivery rather than a standalone modality. Without this reclassification, overlaps arise, such as between digital and psychological interventions (e.g., virtual therapy) or digital and exercise interventions (e.g., wearable activity trackers). Future reviews should explicitly address modality overlaps and classify digital platforms as delivery mechanisms rather than modalities.^46–48^

Another limitation was the predominance of cancer patients in the literature, many of whom underwent neoadjuvant chemotherapy prior to prehabilitation. This sequencing complicates interpretation, as improvements in postoperative outcomes may reflect the effects of neoadjuvant therapy rather than prehabilitation itself. Furthermore, cancer-specific outcomes were not frequently reported and, therefore, excluded from the analysis.^49,50^ Future research is needed to clarify the role of prehabilitation specifically in oncology populations.

Finally, as with all systematic reviews, the possibility of missed studies cannot be excluded. Multiple search strategies and keyword combinations were employed, yet some earlier studies may not have been captured if not indexed with “prehabilitation” as a keyword. Although the likelihood of missing recent publications is low given our utilization of broad search terms, this limitation underscores the importance of ongoing updates as the field evolves.

## Conclusion

Prehabilitation represents a promising and evolving domain within perioperative medicine, as most of the current evidence supports its association with improved surgical outcomes. This review highlights a notable underutilization of prehabilitation strategies across certain surgical specialties, indicating important avenues for future investigation. Furthermore, most existing studies employ single- or dual-modality interventions, underscoring the need for rigorous research into comprehensive, multidisciplinary prehabilitation programs. Such studies should aim to evaluate both short- and long-term outcomes, including patient-centered metrics such as quality of life and mental health.

## Abbreviations

CABG: Coronary Artery Bypass Graft
ENT: Ear, Nose, and Throat
ERAS: Enhanced Recovery After Surgery

## Ethics approval and informed consent

Not applicable

## Consent for publication

Not applicable

## Author’s contributions

EAS analyzed rehabilitation data, conducted comparative analyses across surgical specialties, generated figure visualizations, and contributed substantially to writing and revision. AH assisted with manuscript revisions, gathered updated datasets, and developed supplementary files. DDB proposed critical modifications to strengthen analytical power and address statistical limitations, provided extensive writing and revision, and guided the analytical execution of the manuscript. CJ, SK, and DM contributed to the initial phases of data collection, statistical analyses, figure preparation, and literature review. DCA, KG, and NS designed the research, assigned tasks, led data interpretation, supervised project progress, and contributed significantly to writing and revision. All authors have read and approved the final manuscript, agreed on the target journal, and take full responsibility and accountability for the integrity and content of the article.

## Data Availability

All data necessary to reproduce the findings, including manuscript identifiers and analytical details, are provided in the supplementary table.

## Acknowledgements

Not applicable.

## Collaborating authors or Study Group members

Not applicable.

## Supplementary Table Legends

**Supplementary Table 1.** Comprehensive metadata and manuscript-level details for 153 studies included after application of exclusion criteria. Data are organized by year of publication, surgical specialty, type of prehabilitation modality, study design, reported outcomes, and study conclusions.

